# Global, Regional, and National Burden of Lung Cancer from 1990 to 2021, and Predictions for 2040: Insights from the 2021 Global Burden of Disease Study

**DOI:** 10.1101/2025.08.31.25334803

**Authors:** Dongjing Ma, Shenwen He, Yingying Yang, Keqin Gao, Jinbin Wang, Xingmin Wei, Jianjun Wu

**Affiliations:** School of Public Health, Gansu University of Chinese Medicine, Lanzhou, 730000, Gansu, China

**Keywords:** Lung cancer, Global burden of disease, Incidence, Prevalence, BAPC

## Abstract

**Background:** Lung cancer is the leading cause of cancer-related deaths worldwide and is one of the major diseases contributing to the loss of healthy life years. A comprehensive understanding of the burden of lung cancer is crucial for developing effective prevention and treatment strategies. However, there is still a limited comprehensive assessment of the global burden of lung cancer. This study provides the latest evaluation of lung cancer prevalence, incidence, mortality, and Disability-Adjusted Life Years (DALYs) from 1990 to 2021, systematically analyzes global lung cancer trends from 1990 to 2021, and predicts future trends for 2040, aiming to provide scientific evidence for policy-making.

**Methods:** The data for this study were obtained from the 2021 Global Burden of Disease Study (GBD). The estimated annual percentage changes (EAPCs) were calculated to quantify temporal patterns and assess trends in age-standardized lung cancer prevalence (ASPR), incidence (ASIR), mortality (ASDR), and DALYs; the study conducted stratified analyses by sex, twenty age categories, twenty-one GBD regions, two hundred four countries/regions, and five SDI quintiles; trends were predicted to 2040 using a Bayesian age-period-cohort model. Statistical analyses and plotting were performed using R version 4.4.3.

**Results:** In 2021, the total number of lung cancer cases worldwide was approximately 3,253,729. The global ASPR was 37.3 per 100,000, the ASIR was 26.4 per 100,000, and the ASDR was 23.5 per 100,000 (95% UI: 21.2-25.8), with an age-standardized DALY rate of 533 years lost per 100,000. Regionally, higher socio-demographic index (SDI) areas had the highest ASPR, ASIR, ASDR, and age-standardized DALY rates, while lower SDI areas had relatively lower incidence rates. Geographically, the High-income Asia Pacific region had the highest ASPR, East Asia ranked first in ASIR and ASDR, and Central Europe had the highest age-standardized DALY rate. Among countries, Monaco had the highest ASPR, ASIR, and ASDR, while Greenland had the highest age-standardized DALY rate.

**Conclusion:** This study describes the current global prevalence of lung cancer, estimates the influencing factors in different regions, utilizes scientific statistical methods to predict future trends in lung cancer development, and proposes response measures tailored to the burden of lung cancer in different regions, which will help alleviate the global burden of lung cancer.

## Introduction

Lung cancer is one of the leading causes of cancer-related deaths globally, with significant disparities in disease burden across regions, genders, and socioeconomic levels. According to GLOBOCAN 2020 data, there were approximately 2.207 million new cases of lung cancer worldwide, accounting for 11.4% of all cancers, with 1.796 million deaths, making it the primary reason for cancer-related deaths [1]. The five-year survival rate for lung cancer shows significant variation globally, ranging from 32.9% in Japan to 10%−20% in most countries, with an overall survival rate still below 20%, which presents a major public health challenge [2].

The incidence and mortality rates of lung cancer exhibit marked regional imbalances. In Asia, the age-standardized incidence rate in Taiwan increased from 22.53 per 100,000 in 1994 to 34.09 per 100,000 in 2013 [3]. In South Korea, the lung cancer incidence rate among women has risen at an annual growth rate of 1.7%. Although some regions in Asia have lower lung cancer incidence rates, their mortality rates remain the highest, which is associated with an increase in adenocarcinoma proportions and a rising proportion of adenocarcinomas, which increased from 29.1% in 1999 to 46.7% in 2012 and late-stage diagnoses [4].Data from the Global Burden of Disease (GBD) from 2010 to 2019 indicate that while age-standardized incidence rates in high SDI countries have experienced an average annual decline of 1.2%, middle- and low-SDI countries have seen an increase in incidence rates by 0.8% per year due to tobacco prevalence and delayed tobacco control policies [5]. The World Health Organization (WHO) Global Tobacco Epidemic Report states that if the MPOWER tobacco control policies are fully implemented, over one million lung cancer deaths could be avoided by 2030 [6]. In addition to smoking, air pollution and occupational carcinogens such as arsenic, nickel, radon, asbestos, X-rays, and gamma radiation are also acknowledged as significant risk factors for lung cancer[7]. The distribution of lung cancer incidence rates between men and women shows significant differences, with men generally having higher rates, although it is gradually rising among women [3]. Particularly in high-income countries, the rising smoking rates among women contribute to the increasing trend of lung cancer incidence [8]. The rising incidence of lung cancer across countries and regions is closely tied to shifts in policies, environments, and lifestyle choices [9].

The main objective of this study is to analyze the epidemiological characteristics of lung cancer globally in 2021 and explore the differences across regions, genders, and age groups. We utilize the latest global burden of disease data to detail the changes in lung cancer prevalence, incidence, mortality, Disability-Adjusted Life Years (DALY) rates, and their correlation with SDI from 1990 to 2021. Additionally, by analyzing the changes in lung cancer burden across regions, we identify potential social and environmental factors influencing lung cancer incidence, emphasizing the importance of regional disparities. We also highlight the specificity of gender and age differences in lung cancer burden, analyze the proportion of lung cancer burden attributable to related environmental risk factors, and make corresponding predictions about future trends in lung cancer development. This study proposes policy recommendations for lung cancer prevention and control tailored to the characteristics of different regions, which is of significant practical importance for reducing the global burden of lung cancer. In summary, this study aims to provide a scientific basis for the formulation of future lung cancer prevention and control policies through systematic epidemiological analysis, promoting improvements in public health globally.

## Methods

### Data Sources

Lung cancer data from 1990 to 2021 were obtained through the Global Health Data Exchange (GHDx) query tool. The survey included information on prevalence, incidence, mortality, and Disability-Adjusted Life Years (DALYs); age-standardized prevalence rate (ASPR), age-standardized mortality rate (ASDR), age-standardized incidence rate (ASIR), and age-standardized DALY rate (ASR) were sourced from GBD 2021. The research methods used in GBD have been widely described in existing literature. The SDI index quantifies the level of development of a country or region using fertility rates, education levels, and per capita income data [10, 11]. The SDI ranges from 0 to 1, with higher values indicating better socioeconomic development. It is known that SDI is associated with disease incidence and mortality rates. In this study, we categorized countries and regions into low, lower-middle, middle, upper-middle, and high SDI categories to investigate the relationship between lung cancer and social development. Risk Factor Assessment: In addition to primary indicators such as prevalence, incidence, mortality, and DALYs, this study also examined the impact of specific risk factors on the burden of lung cancer.

### Four key lung cancer risk factors were emphasized

Tobacco, particulate matter pollution, occupational carcinogens, and residential radon. We analyzed lung cancer-related mortality and DALYs attributable to these factors, stratified by region, to elucidate their geographical differences in impact.

### Statistical Analysis

To assess trends in age-standardized rates (ASR) of lung cancer incidence, mortality, DALYs, and prevalence, the study employed estimated annual percentage changes (EAPC). ASR was calculated per 100,000 population using the following formula:

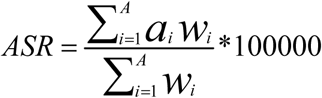

(a_i_: specific age rate for that age group; W_i_: number of individuals corresponding to the age group in the standard population; A: number of individuals in the age group).

EAPCs were calculated based on a regression model that characterizes the pattern of age-standardized rates over specific periods [12]. The formula is: Y=α+βX+e, where Y is the natural logarithm of ASR, X is the year, α is the intercept, β is the slope or trend, and e is the error term. EAPC is calculated as 100×[exp(β) - 1], representing the annual percentage change. A linear regression model was used to calculate the 95% confidence interval (CI) for EAPC. If the EAPC and its 95% CI lower limit are positive, the ASR is considered to be increasing. Conversely, if the EAPC and its 95% CI upper limit are negative, the ASR is considered to be decreasing. If neither condition is met, the age-standardized rate is deemed stable. Spearman correlation was used to assess the association between SDI and age-standardized lung cancer rates. In this study, we utilized a Bayesian age-period-cohort (BAPC) model combined with integrated nested Laplace approximation to predict future trends in lung cancer burden, as previous studies have shown that BAPC offers better coverage and accuracy compared to alternative predictions [13–16]. The computation process used the R language BAPC package [16].

## Results

### Global Level

In 2021, the global burden of lung cancer was severe, with approximately 3,253,729 cases (95% UI: 2,947,830.8-3,558,836.1), a decrease of 232.54% from 1990. The absolute number of cases significantly declined, with the ASPR showing an increase from 34.3 per 100,000 in 1990 (95% UI: 32.7-35.7) to 37.3 per 100,000 in 2021 (95% UI: 33.8-40.8). The EAPC for ASPR was 0.4 (95% CI: 0.27 - 0.52) (Table 1, Figure 1). In 2021, the number of lung cancer cases reached 2,280,688.2 (95% UI: 2,063,251.9-2,509,739.7), a decrease of 201.46% from 1990. The ASIR for lung cancer decreased from 28.5 per 100,000 in 1990 (95% UI: 27.1-29.9) to 26.4 per 100,000 in 2021 (95% UI: −0.31 ∼ −0.17). The EAPC for ASIR was −0.24 (95% CI: −0.31 ∼ −0.17), indicating a decline in lung cancer incidence, although the decrease was limited (Table 1, Figure 1). In 2021, the number of lung cancer deaths was 2,016,547.4 (95% UI: 1,820,497.7-2,218,371.9), with an ASDR of 23.5 per 100,000 (95% UI: 21.2-25.8), and an EAPC of −0.54 (95% CI: −0.6 ∼ −0.48) (Table 1, Figure 1). The global DALYs for lung cancer in 2021 were approximately 46,536,272.1 (95% UI: 41,903,412.3-51,205,051.4), with an age-standardized DALY rate of 533 (95% UI: 480.1-586.4), and an EAPC of −0.87 (95% CI: −0.93 ∼ −0.81) (Table 1, Figure 1).

**Figure 1:**
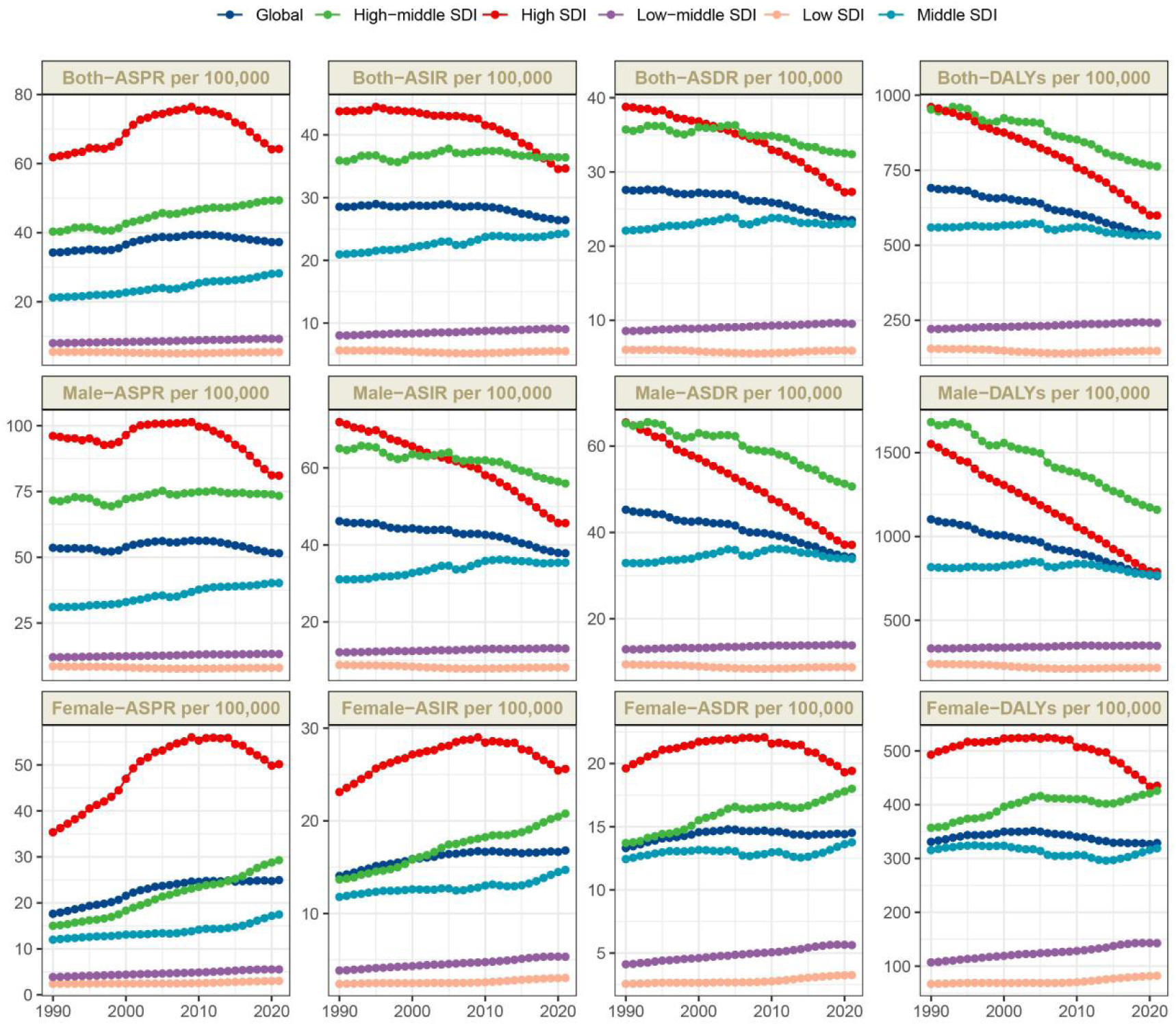
Trends in ASPR, ASIR, ASDR, and DALYs in lung cancer from 1990 to 2021.

**Table. 1:**
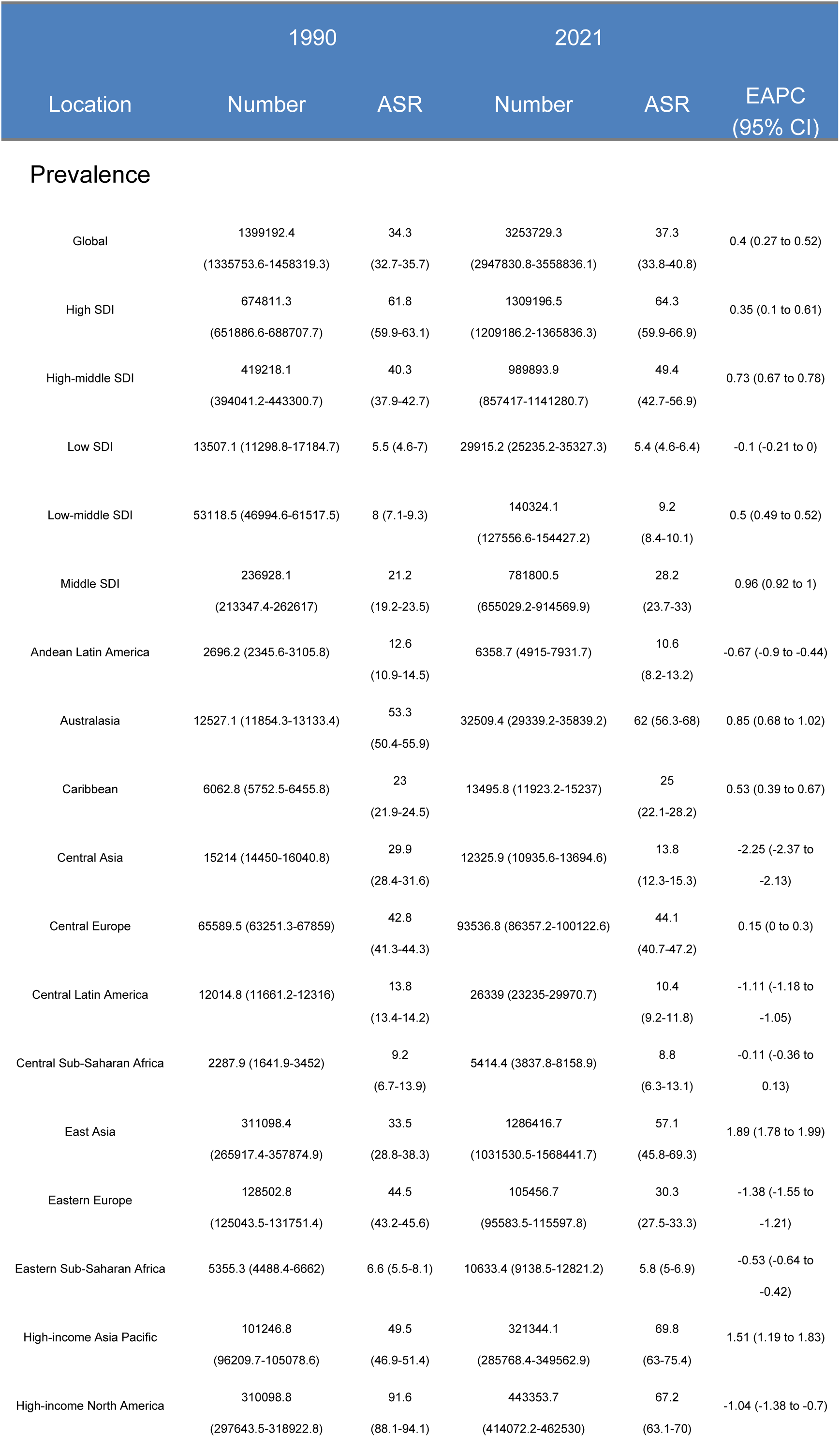
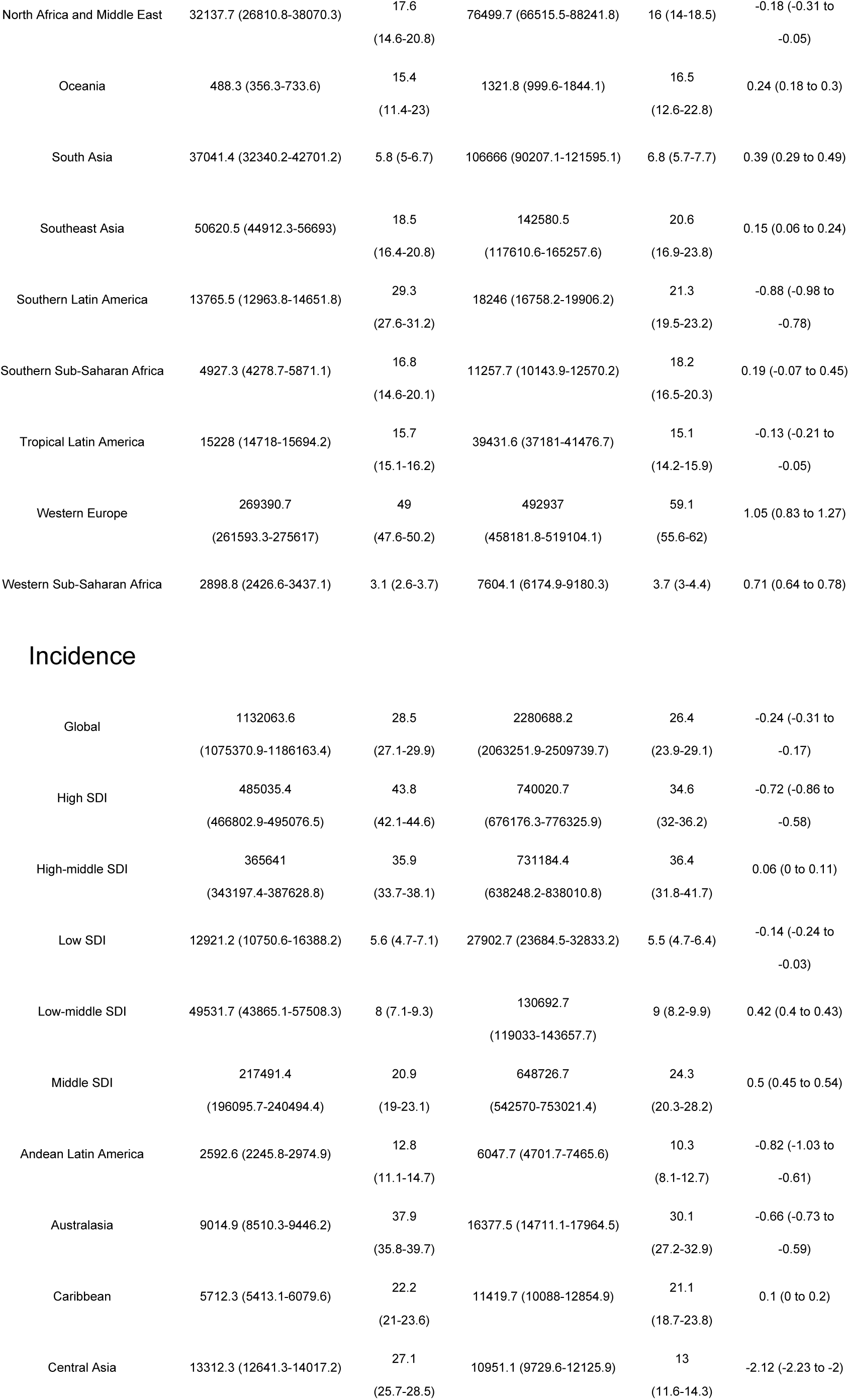
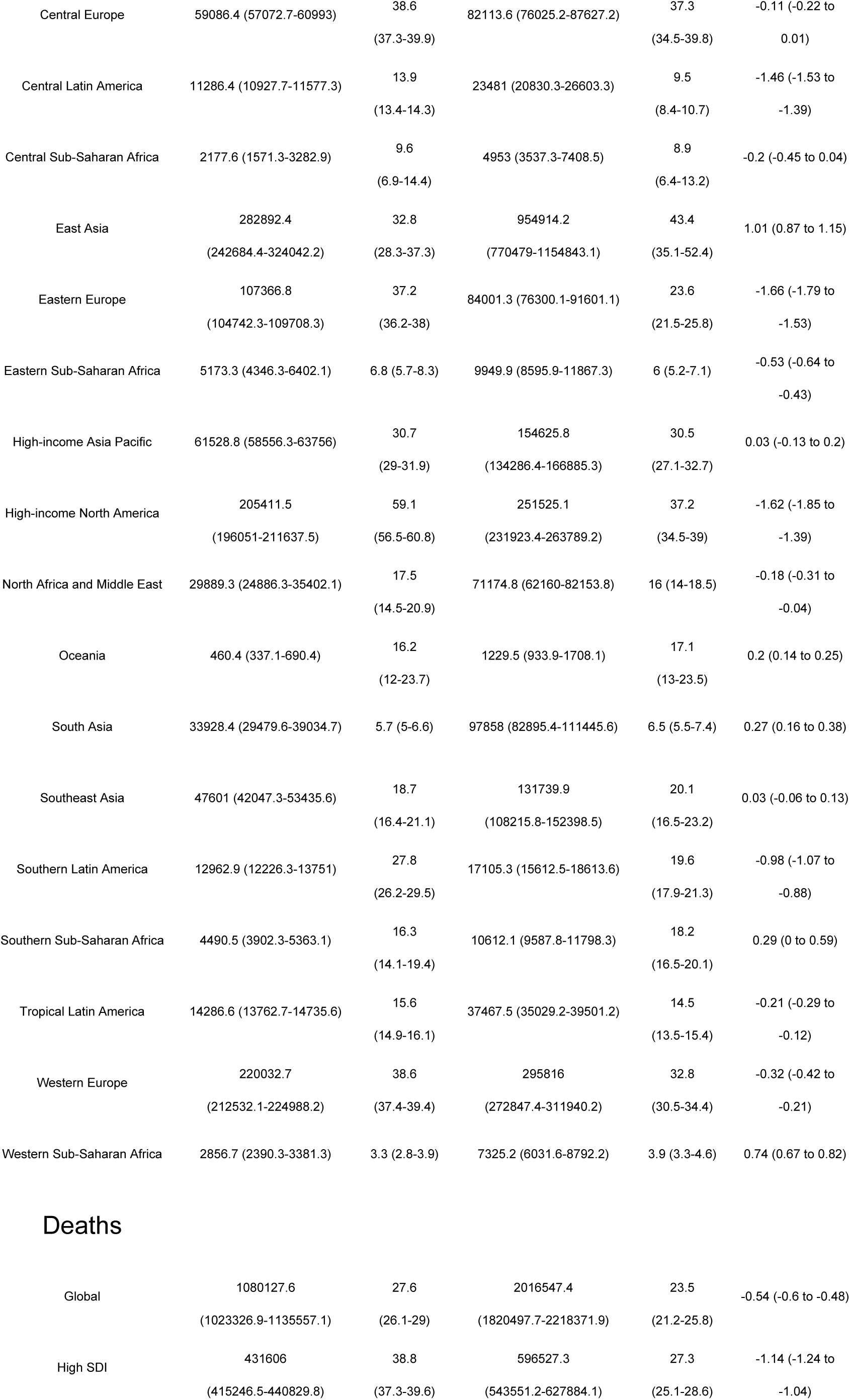
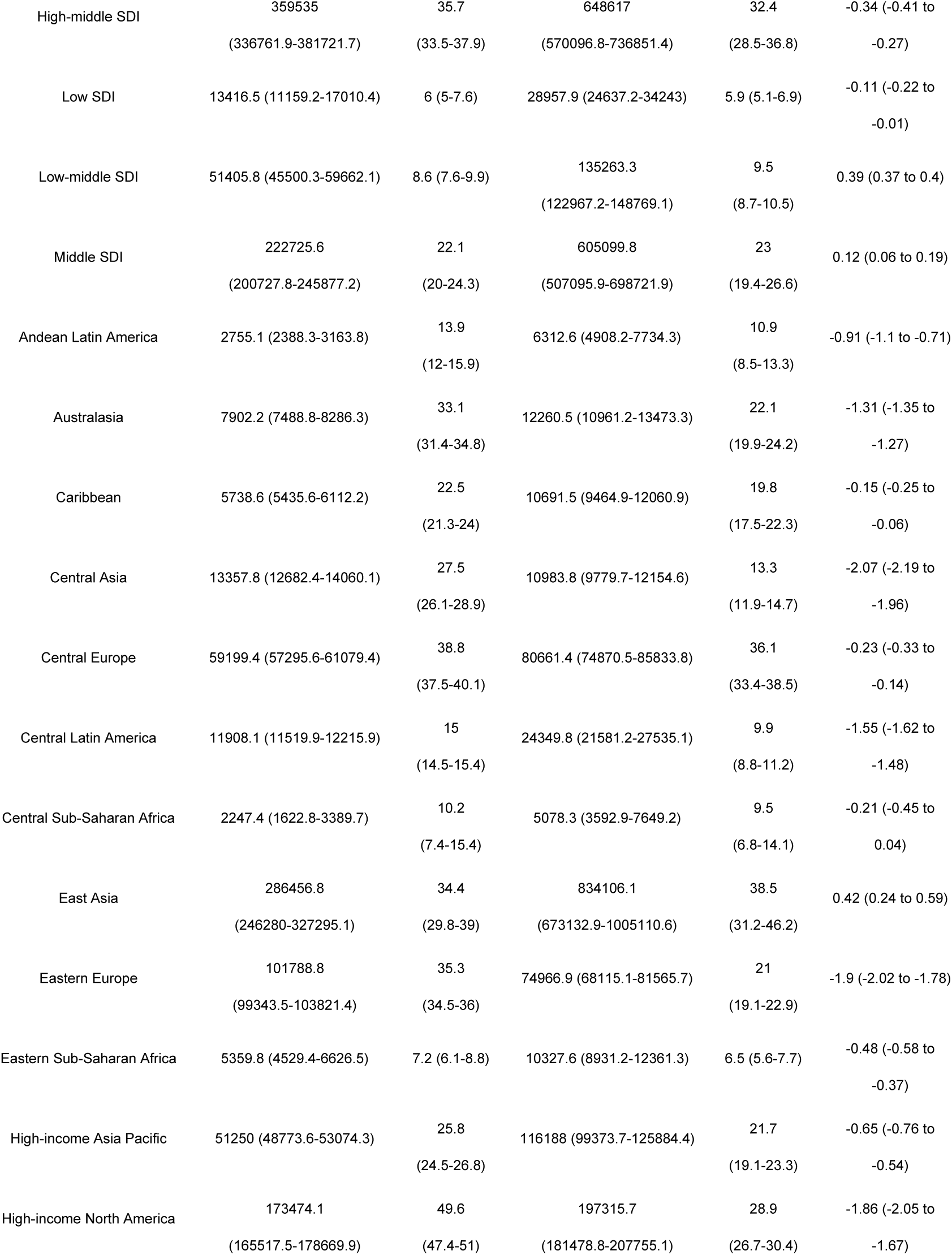
Global and regional trends in lung cancer burden: prevalence, incidence, mortality, and disability-adjusted life years (1990–2021).

### Regional Level

The global burden of lung cancer exhibited significant regional differences closely related to socio-demographic index (SDI) levels. ASPR showed significant disparities, with high SDI regions reporting the highest incidence and prevalence, totaling 1,309,196.5 cases (95% UI: 1,209,186.2-1,365,836.3) and an incidence rate of 64.3 per 100,000 (95% UI: 59.9-66.9), while low SDI regions had the lowest incidence and prevalence, with 29,915.2 cases (95% UI: 25,235.2-35,327.3) and an incidence rate of only 5.4 per 100,000 (95% UI: 4.6-6.4) (Table 1, Figure 1). The temporal trend of ASPR revealed different patterns across SDI levels, potentially indicating different stages of epidemiological transition. High-middle SDI and middle SDI regions both exhibited significant upward trends, with EAPCs of 0.73 (95% CI: 0.67 - 0.78) and 0.96 (95% CI: 0.92 - 1), respectively. In high-middle SDI regions, ASPR increased from 40.3 per 100,000 in 1990 (95% UI: 37.9-42.7) to 49.4 per 100,000 in 2021 (95% UI: 42.7-56.9), while in middle SDI regions, ASPR rose from 21.2 per 100,000 in 1990 (95% UI: 19.2-23.5) to 28.2 per 100,000 in 2021 (95% UI: 23.7-33), indicating an increasing burden of lung cancer in these regions (Table 1, Figure 1). In contrast, only the low SDI region showed a decrease in EAPC, which was only 0.1 (95% CI: −0.21 to 0), failing to demonstrate effective measures taken to reduce the lung cancer burden in these areas (Table 1, Figure 3A). ASIR and ASDR further emphasized these regional differences. In 2021, the highest ASIR and ASDR were reported in middle-high SDI regions, with high SDI regions slightly lower than middle-high SDI regions, and low SDI regions reporting the lowest. The ASIR for middle-high SDI regions was 34.6 per 100,000 (95% UI: 32-36.2), while low SDI regions had an ASIR of 5.5 per 100,000 (95% UI: 4.7-6.4). Similarly, the ASDR for middle-high SDI regions was 32.4 per 100,000 (95% UI: 28.5-36.8), while low SDI regions had an ASDR of 5.9 per 100,000 (95% UI: 5.1-6.9) (Table 1, Figure 1, Figure 2C). This indicates that the higher the SDI region, the heavier the lung cancer burden. The DALY rate for lung cancer in middle-high SDI regions was 762.7 per 100,000 (95% UI: 667.1-870), while the DALY rate for low SDI regions was 147.6 per 100,000 (95% UI: 125.2-174.4) (Table 1, Figure 1, Figure 2D). This suggests that regions with higher SDI levels face severe lung cancer burdens and may lack effective means to reduce lung cancer incidence and address lung cancer prognosis, posing increasing challenges for high SDI and middle-high SDI regions.

**Figure 2:**
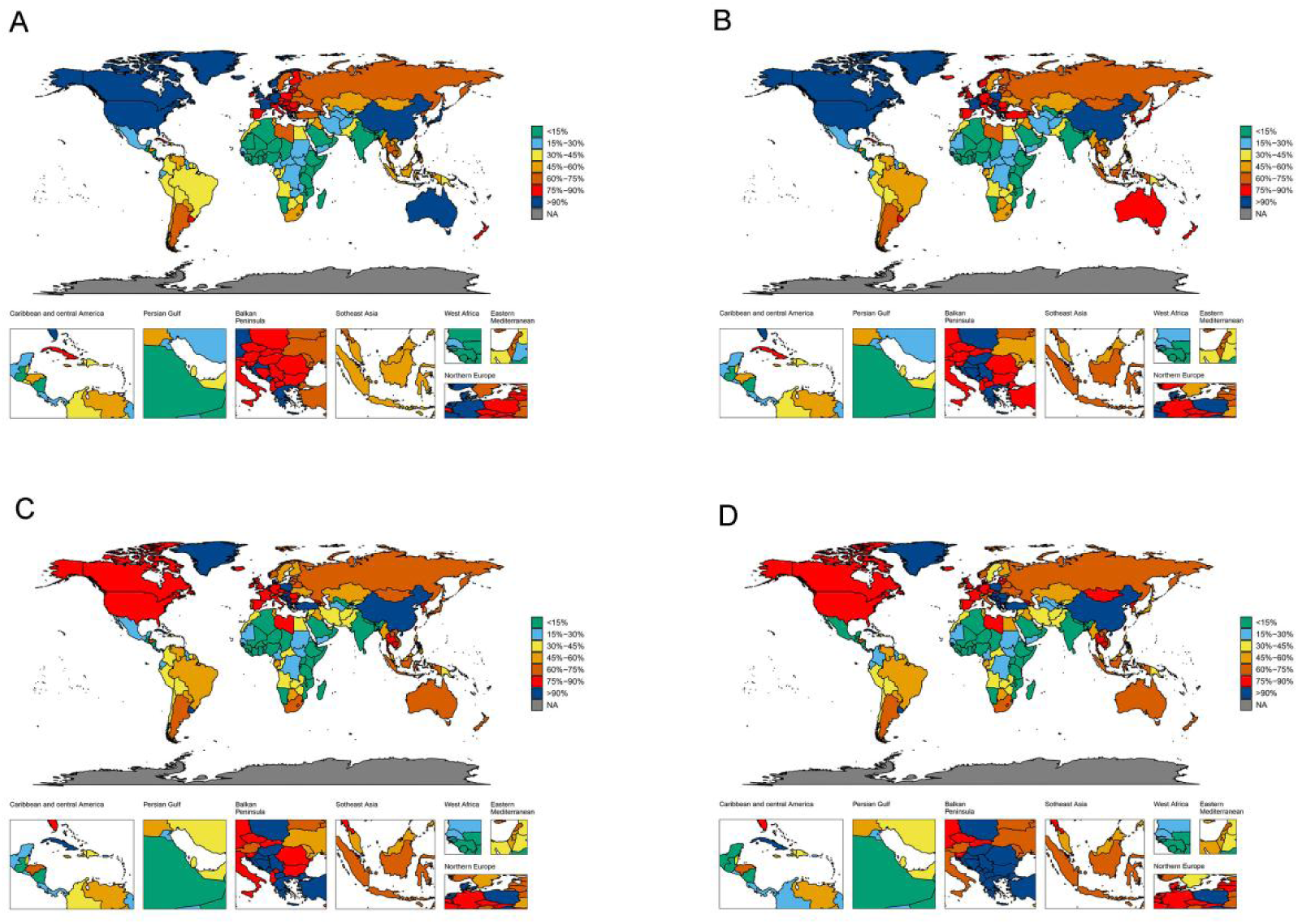
The global disease burden of lung cancer for both sexes in 204 countries and territories. (A) Prevalence rate. (B) Incidence rate. (C) Death rate. (D) DALYs rate.

**Figure 3:**
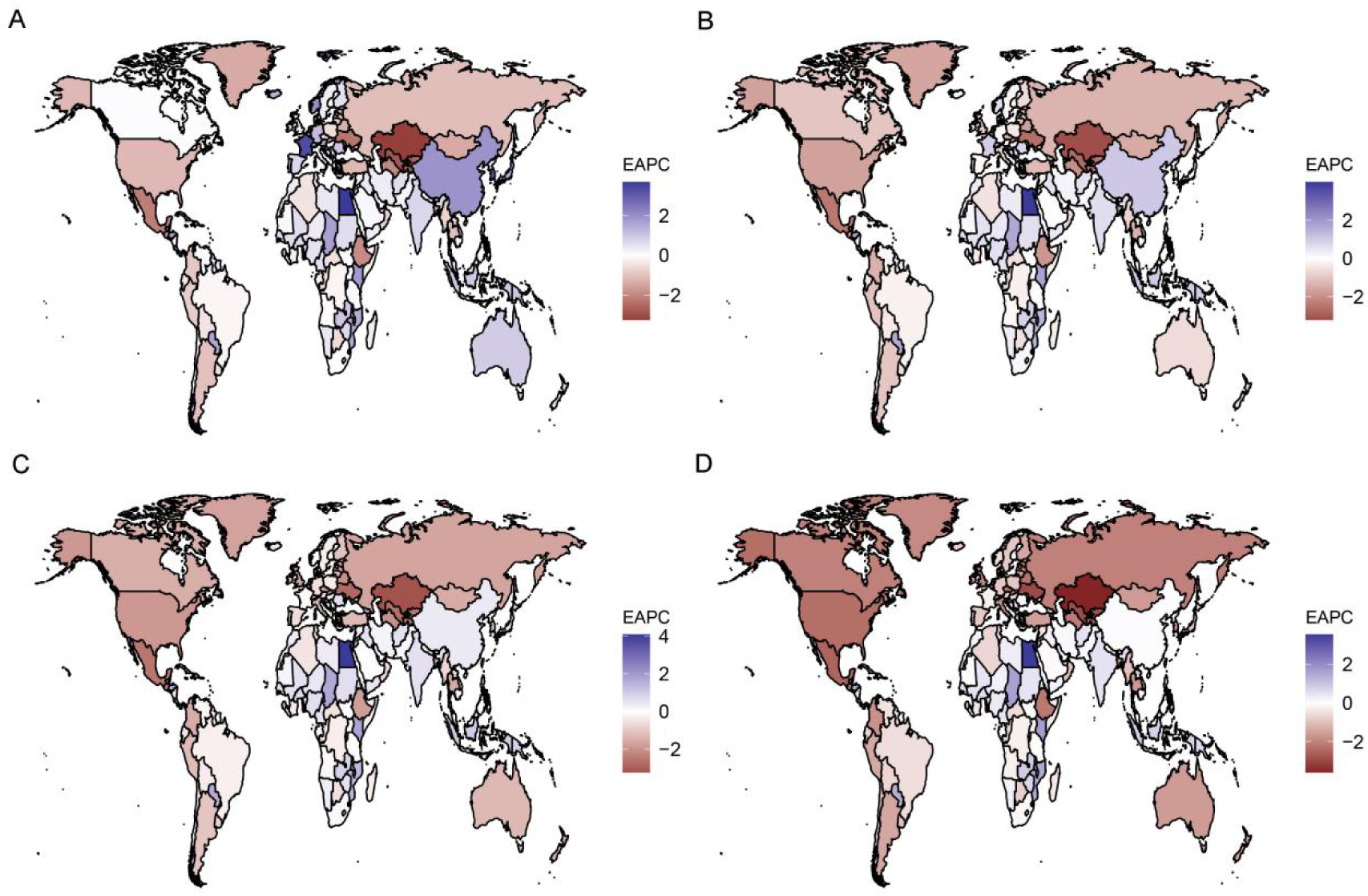
The global disease burden of lung cancer for both sexes in 204 countries and territories. (A) EAPC for prevalence. (B) EAPC for incidence. (C) EAPC for deaths. (D) EAPC for DALYs.

Geographically, the High-income Asia Pacific region had the heaviest lung cancer burden globally, with an ASPR of 69.8 per 100,000 (95% UI: 63-75.4) in 2021, followed closely by High-income North America with an ASPR of 67.2 per 100,000 (95% UI: 63.1-70). Western Europe and East Asia had ASPRs of 59.1 per 100,000 (95% UI: 55.6-62) and 57.1 per 100,000 (95% UI: 45.8-69.3), ranking fourth and fifth among the studied regions (Table 1, Figure 2A). The lung cancer prevalence in high-income regions far exceeds that of low-income regions.

The region with the highest global ASIR is East Asia, reporting an ASIR of 43.4 per 100,000 (95% UI: 35.1-52.4) in 2021. In contrast, Western Sub-Saharan Africa had the lowest ASIR at only 3.9 per 100,000 (95% UI: 3.3-4.6) (Table 1, Figure 2B). Furthermore, the time trends from 1990 to 2021 indicate different patterns within Asia. East Asia exhibited the most significant increase in ASIR, with an EAPC of 1.01 (95% CI: 0.87 to 1.15). Conversely, Central Asia showed the most significant decline, with an EAPC of −2.12 (95% CI: −2.23 to −2) (Table 1, Figure 3B). These starkly different trends highlight the complexity and dynamic changes in lung cancer incidence across different Asian regions. East Asia and Central Europe reported extremely high ASDRs, with East Asia’s ASIR at 38.5 per 100,000 (95% UI: 31.2-46.2) and Central Europe’s ASIR at 36.1 per 100,000 (95% UI: 33.4-38.5), indicating a need for special attention due to high incidence and mortality rates. From 1990 to 2021, ASDRs for lung cancer declined in most regions, with increases only observed in Western Sub-Saharan Africa (EAPC: 0.77, 95% CI: 0.69 to 0.84), East Asia (EAPC: 0.42, 95% CI: 0.24 to 0.59), Oceania, South Asia, and Southern Sub-Saharan Africa, with the frequency of increases being low. Meanwhile, most regions showed a declining trend, with Central Asia experiencing the most significant decline (EAPC: −2.07, 95% CI: −2.19 to −1.96) (Table 1, Figure 3C). Our results indicate that the region with the highest age-standardized DALY rate is Central Europe, with a loss of 896.3 years per 100,000 (95% UI: 830.5-955.3). East Asia ranks second, with a loss of 869.5 years per 100,000 (95% UI: 702.9-1050.6) (Table 1, Figure 2D). From 1990 to 2021, the age-standardized DALY rate for lung cancer increased the most in Western Sub-Saharan Africa (EAPC 0.61, 95% CI: 0.55 to 0.68) and decreased the most in Central Asia (EAPC −2.5, 95% CI: −2.59 to −2.41) (Table 1, Figure 3D). It is noteworthy that DALY rates are declining in most regions, indicating progress in global lung cancer treatment levels. Additionally, the age-standardized DALY rate is negatively correlated with SDI (Supplementary Figure S1).It is also observed that in regions with higher socioeconomic development index (SDI), the proportion of lung cancer cases among younger individuals is lower, while in regions with higher socioeconomic development index (SDI) in 1990 and 2021, the proportion of lung cancer cases among older individuals is higher, primarily concentrated in the 65-74 age group (Figure 4).

**Figure 4:**
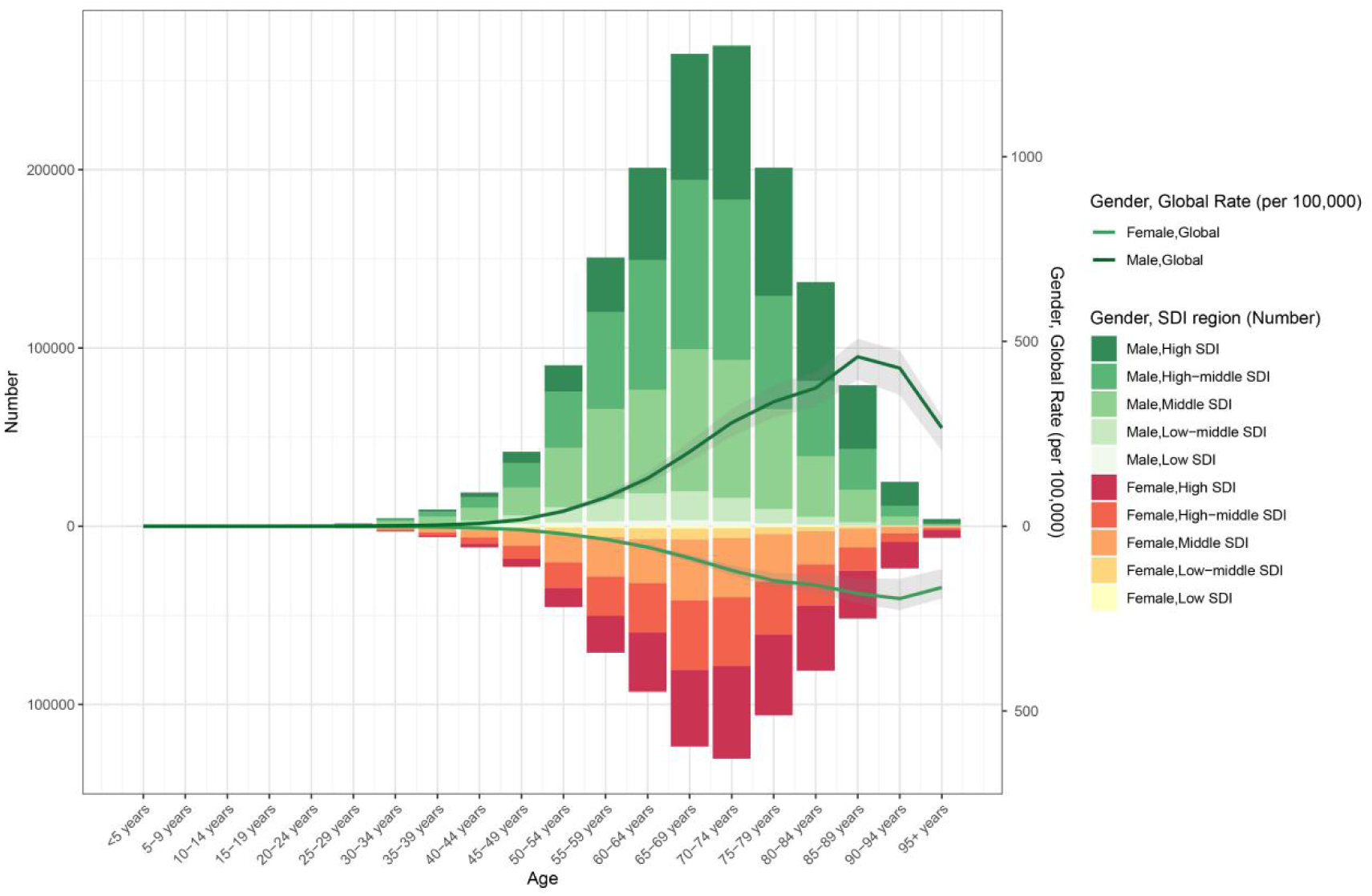
The age-specific numbers and ASIRs of lung cancer by SDI regions in 2021.

### National level

Among all countries, the three countries with the highest ASPR are: Monaco (123.5 per 100,000; 95% UI: 98.8-156), France (89.3 per 100,000; 95% UI: 80.5-98), and Japan (71.2 per 100,000; 95% UI: 64.4-76.5) (Figure 2A, Supplementary Table S1). Notably, two of the three countries with the highest ASPR are located in Western Europe, while Japan is in the High-income Asia Pacific region. This finding aligns with our regional analysis, indicating that both the High-income Asia Pacific and Western Europe are major areas affected by lung cancer. The consistency between national and regional data emphasizes the need for focused attention on the lung cancer burden in economically developed areas. The most significant decline in lung cancer ASPR was observed in Egypt (EAPC of 3.68, 95% CI of 3.19 to 4.17), followed by France (EAPC of 89.3, 95% CI of 80.5-98) (Figure 3A, Supplementary Table S1).

Monaco had the highest ASIR (76.9 per 100,000; 95% UI: 62.1-94.9), followed by Greenland (63.3 per 100,000; 95% UI: 51.1-77.4). In Egypt, there was a wide range of incidence rates and a significant increase in prevalence (EAPC of 3.99, 95% CI: 3.44 to 4.53) (Figure 3B, Supplementary Table S1). The differences in incidence rates among countries within the same region suggest significant variations in risk factors, healthcare systems, and prevention strategies. In 2021, the country with the highest ASDR was Monaco (64.1 per 100,000; 95% UI: 52.2-78.6), with an EAPC of 0.92 (95% CI: 0.56 to 1.28), followed by Greenland (63.9 per 100,000; 95% UI: 51-77.7), with an EAPC of −1.7 (95% CI: −1.83 to −1.56). Monaco’s high mortality rate is rising (Supplementary Table S1). The highest DALYs were reported in Greenland (1525.2 per 100,000; 95% UI: 1236.5-1849.9), followed closely by Monaco (1523.7 per 100,000; 95% UI: 1233.6-1890.2). It is noteworthy that Monaco ranks high in ASPR, ASIR, and ASDR, indicating that these countries face particularly high lung cancer burdens, affecting their population’s mortality and overall health-related quality of life. Egypt (EAPC: 4.09, 95% CI: 3.53 to 4.66) and Lesotho (EAPC: 2.82, 95% CI: 2.47 to 3.17) showed the highest increases in mortality (Figure 3C and Supplementary Table S1), while Egypt (EAPC: 3.54, 95% CI: 3.05 to 4.04) and Lesotho (EAPC: 3.06, 95% CI: 2.65 to 3.47) had the highest years of life lost from age-standardized DALYs from lung cancer. The consistency across all four lung cancer indicators suggests that these countries should place greater emphasis on prevention and treatment strategies for lung cancer.

### Age and Gender Patterns

In 2021, the global ASPR for lung cancer gradually increased with age, peaking at 75-79 years (Figure 5). The study found that the ASPR among men in the same age group consistently exceeded that of women, although the ASPR for women is rising rapidly in both global and higher SDI regions. Notably, the ASPR for men aged 40 to 69 in 2021 showed a slight decline compared to 1990 (Figure 5). The ASIR for lung cancer also gradually increased with age, peaking among individuals aged 85-89 (Supplementary Figure S2). Compared to 1990, the ASDR for lung cancer decreased for both men and women in 2021, but continued to rise with age, with men generally having higher ASDRs than women across most age groups (Supplementary Figure S3). Similarly, DALYs showed a decreasing trend in 2021 compared to 1990, peaking among individuals aged 85-89 (Supplementary Figure S4). This indicates that with advancements in medical care, there has been a certain degree of reduction in lung cancer mortality and DALYs lost.

**Figure 5:**
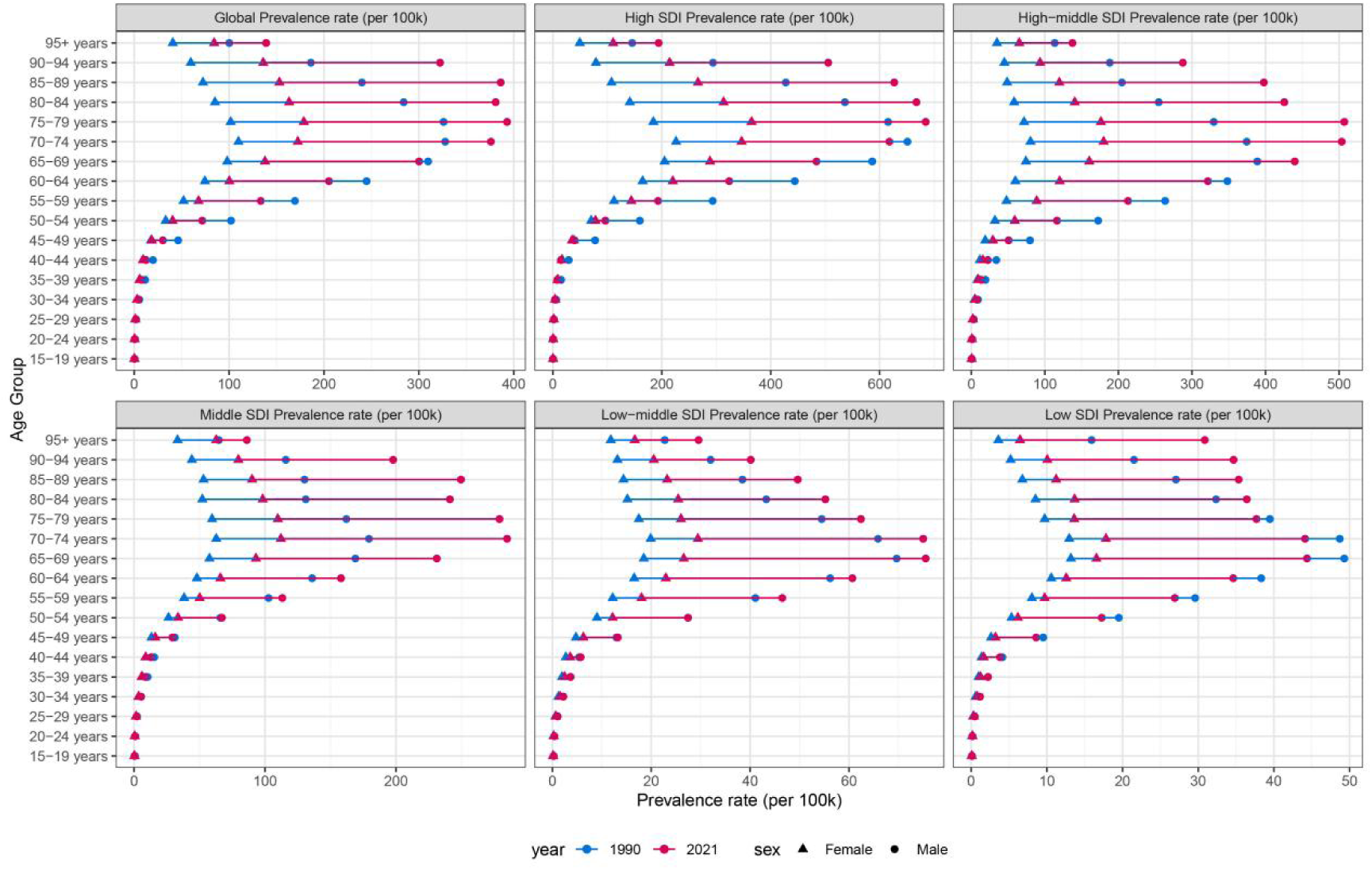
Age-standardized prevalence rates of lung cancer by sex, age group, and socio-demographic index, 1990 and 2021.

### Risk Factors for Lung Cancer

This study collected data on lung cancer-related deaths and DALYs attributable to four risk factors: Tobacco, Particulate matter pollution, Occupational carcinogens, and Residential radon, categorized according to GBD 2021 and further stratified by region (Figure 6). Globally, tobacco was the most significant factor contributing to lung cancer deaths and DALYs in 2021, accounting for 61.4% of all deaths. Regionally, Eastern Europe had the highest proportion of deaths attributable to tobacco at 66.7%, followed closely by East Asia at 66.6%. Western Sub-Saharan Africa had the lowest at 25.7%. Particulate matter pollution also played a critical role, with Eastern Sub-Saharan Africa having the highest proportion at 40%, while High-income North America had the lowest at 3.3%. Occupational carcinogens had a significant impact, with Australasia reporting the highest proportion at 33.4%, and Oceania the lowest at 6.4%. Residential radon contributed to deaths to some extent, with Central Asia having the highest proportion at 7.4%, and Australasia the lowest at 1.4%. The impact of risk factors on mortality varied across different SDI regions, with lower SDI regions experiencing slightly lower tobacco-related deaths compared to higher SDI regions, while particulate matter pollution caused significantly higher deaths in lower SDI regions compared to higher SDI regions. Globally, tobacco use remains the primary risk factor for lung cancer, accounting for 61.8% of all DALYs. Regionally, Central Europe had the highest proportion of tobacco-related DALYs at 68.7%, while Eastern Sub-Saharan Africa had the lowest at 24.8%. Particulate matter pollution also significantly impacted DALYs, with Eastern Sub-Saharan Africa having the highest proportion at 39.6%, and High-income North America the lowest at 3.3%.Occupational carcinogens also had a considerable impact on DALYs, with Australasia reporting the highest proportion at 29.4%, and Oceania the lowest at 6.6%. Residential radon also played a role in DALYs, with Central Asia having the highest proportion at 7.3%, and Australasia the lowest at 1.5%. For the five SDI regions, the impact of risk factors on DALYs was generally consistent with their impact on mortality, highlighting the complex interactions of these factors in the global burden of lung cancer.

**Figure 6:**
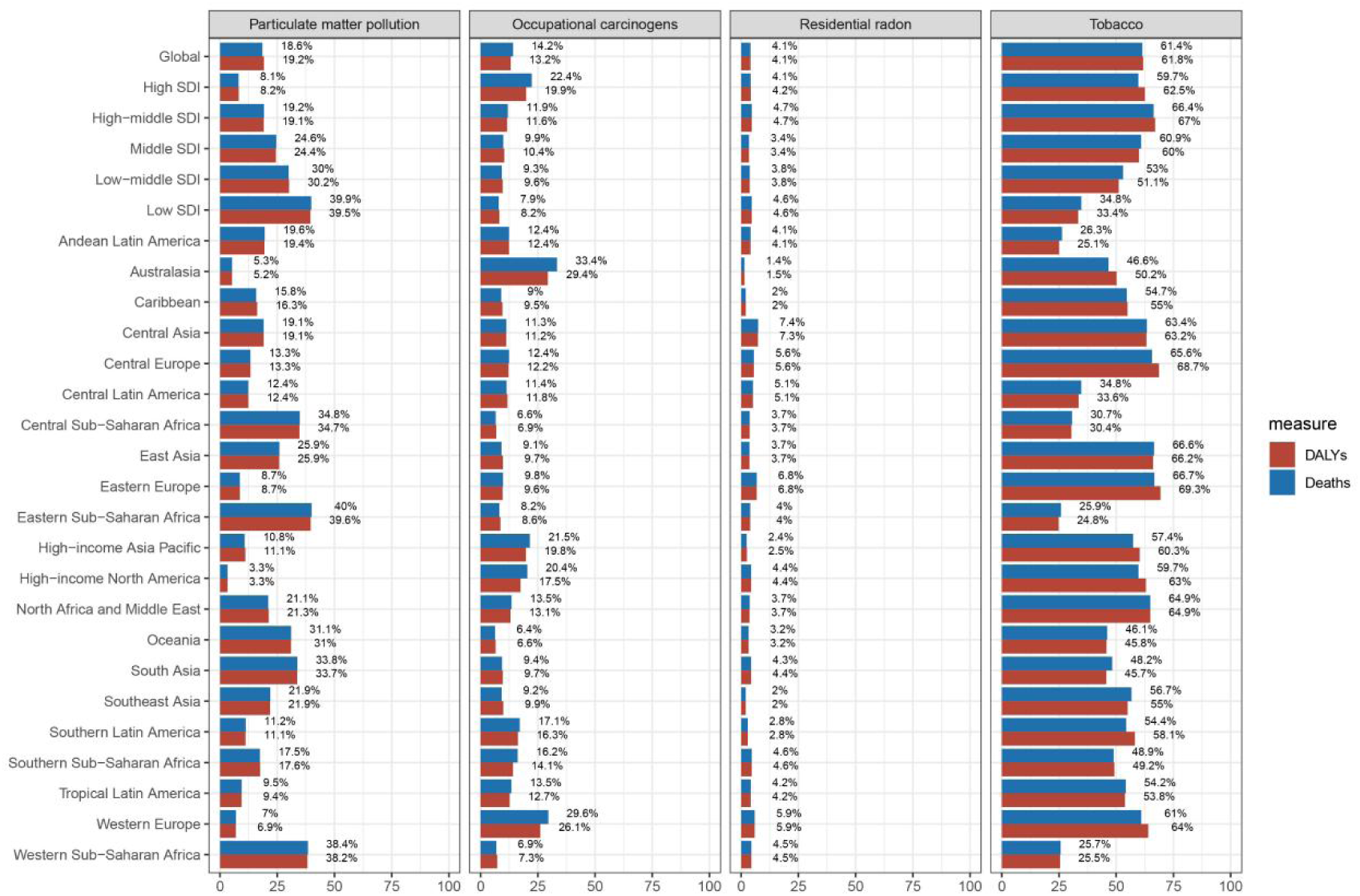
Percentage of age-standardized DALYs rates and ASDR of lung cancer attributable to Tobacco、Particulate matter pollution、Occupational carcinogens and Residential radon.

### Future Predictions for the Global Lung Cancer Burden

It is expected that from 2021 to 2040, the global burden of lung cancer will undergo significant changes. Figure 7 illustrates the future predictions for lung cancer based on GBD. The global ASPR for lung cancer is projected to decline to some extent. By 2040, the ASPR for lung cancer is expected to decrease to 31.31 per 100,000, a reduction of 16.01% from 2021. The ASPR for men is projected to be 40.83 per 100,000, a decrease of 20.72% from 2021; for women, the ASPR is expected to be 23.52 per 100,000, a decrease of 5.8% from 2021. The ASIR for lung cancer is also estimated to decline, from approximately 34.69 per 100,000 in 2021 to about 26.93 per 100,000 in 2040, with men decreasing from approximately 37.84 per 100,000 in 2021 to about 29.77 per 100,000 in 2040, and women decreasing from approximately 32.4 per 100,000 in 2021 to about 24.8 per 100,000 in 2040. Over time, both male and female incidence and prevalence rates are gradually declining, indicating that as society develops, we will further understand the causes of lung cancer and implement scientific prevention and management measures. The predicted ASDR for lung cancer shows that global lung cancer deaths will decrease from approximately 23.05 per 100,000 in 2021 to about 22.45 per 100,000 in 2040, with men decreasing from approximately 31.19 per 100,000 in 2021 to about 28.2 per 100,000 in 2040, and women increasing from approximately 15.12 per 100,000 in 2021 to about 17.39 per 100,000 in 2040. This indicates that the decline in lung cancer mortality is limited, and the treatment situation remains pessimistic, posing a significant ongoing threat of mortality. We also estimate that the age-standardized DALY rate for lung cancer will continue to decline, from 533 years lost per 100,000 in 2021 to 444.36 years in 2040. Among these four indicators, men consistently have higher rates than women, but the gap is gradually narrowing over time.

**Figure 7:**
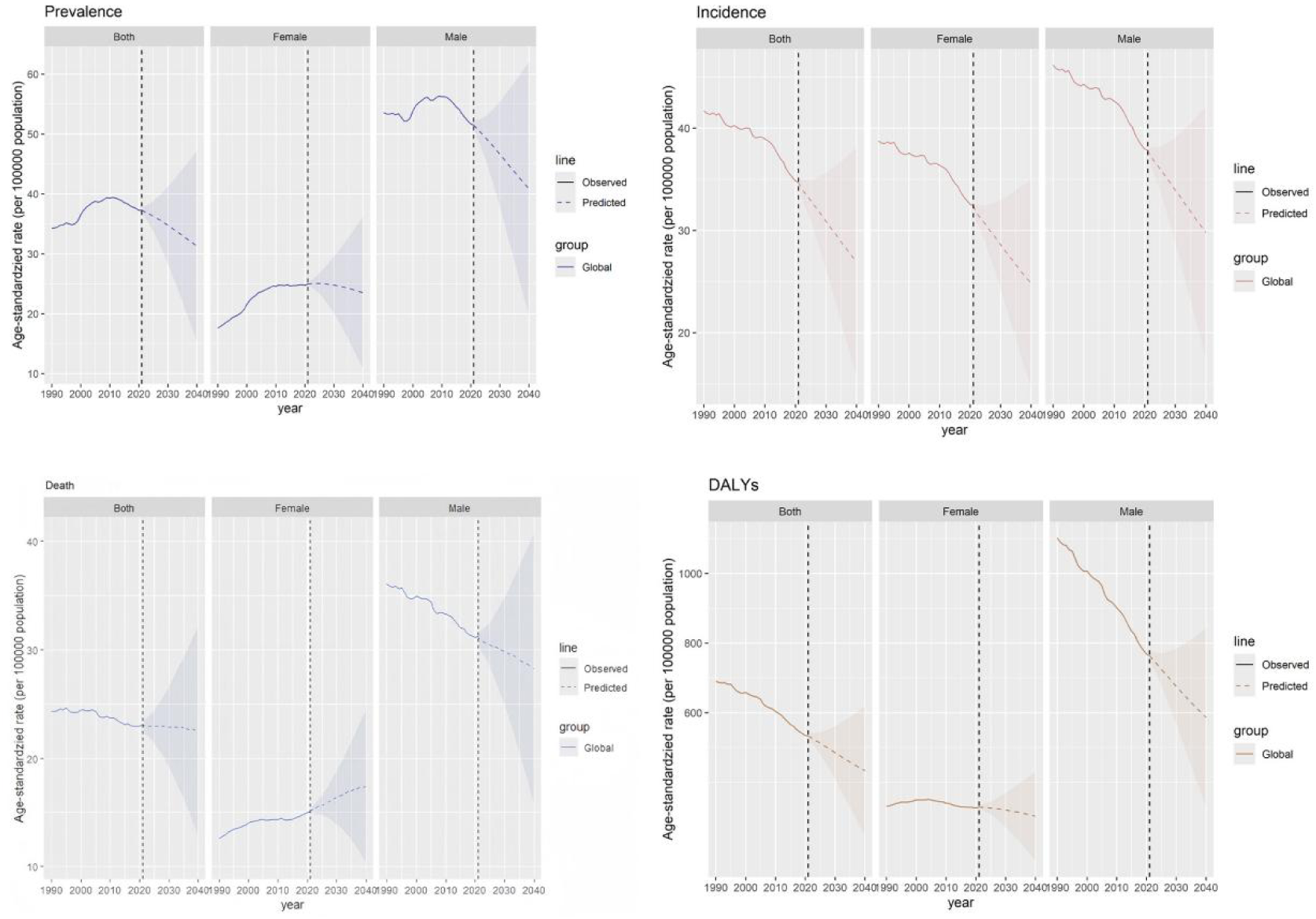
Future forecasts of global burden of lung cancer. Prevalence is ASPR, Incidence is ASIR, Death is ASDR, and DALYs is age-standardized DALY rates.

## Discussion

This study systematically assessed the epidemiological characteristics of lung cancer globally from 1990 to 2021 based on the 2021 Global Burden of Disease (GBD) data and its association with the Socio-demographic Index (SDI). The results indicate that although the global age-standardized incidence rate (ASIR) and mortality rate (ASDR) for lung cancer show a slow declining trend (ASIR EAPC: −0.24; ASDR EAPC: −0.54), the absolute number of cases and deaths has significantly increased, with 2.281 million new lung cancer cases and 2.017 million deaths reported globally in 2021, representing increases of 201.46% and 86.7%, respectively, compared to 1990. This paradoxical phenomenon is primarily attributed to population growth and aging, particularly in low- and middle-income countries (LMICs), where the demographic transition overlaps with the burden of non-communicable diseases, exacerbating the absolute burden of lung cancer [17].

From 1990 to 2021, the estimated annual percentage change (EAPC) for the global lung cancer age-standardized prevalence rate (ASPR) was 0.4 (95% CI: 0.27–0.52), indicating a slight increase in ASPR. Although the EAPC for ASIR was −0.24 (95% CI: −0.31–−0.17), suggesting a decline in incidence, the absolute number of cases increased by 201.46%, reflecting the combined effects of population growth and aging. This phenomenon aligns with the epidemiological transition of global non-communicable diseases (NCDs), where, alongside economic development, the burden of infectious diseases declines while the burden of chronic diseases like lung cancer rises [18]. Notably, high SDI regions (such as High-income Asia Pacific) reported an ASPR as high as 69.8 per 100,000, significantly exceeding that of low SDI regions (5.4 per 100,000), closely linked to historically high smoking rates, exposure to air pollution, and improved diagnostic capabilities in high-income countries [19]. In terms of mortality, the global ASDR EAPC was −0.54 (95% CI: −0.6–−0.48), but mortality rates in East Asia (EAPC=0.42) and parts of Sub-Saharan Africa (such as West Africa, EAPC=0.77) are still rising. The high mortality rate in East Asia may be attributed to worsening PM2.5 pollution due to rapid urbanization [20], while the situation in Africa is related to a lack of medical resources and a high proportion of late-stage diagnoses. Additionally, disruptions in healthcare systems during the COVID-19 pandemic may have exacerbated delays in lung cancer diagnosis and treatment in low- and middle-income countries [21], but further research is needed to verify this.

The relationship between SDI and lung cancer burden is nonlinear. High SDI regions (such as Monaco and Japan) rank among the highest globally for ASPR, ASIR, and ASDR, while low SDI countries (such as Burkina Faso) have the lowest disease burden. This seemingly paradoxical phenomenon reflects differences in exposure across various stages of development and the complex interactions of multiple factors. For instance, high SDI regions have experienced long-term tobacco epidemics and industrial pollution accumulation [22]. During economic transitions, the increase in female smoking rates due to social liberation, greater air pollution, improved diagnostic capabilities leading to higher detection rates, and the rising exposure to risk factors associated with rapid urbanization and lifestyle changes have all contributed to the increased lung cancer burden in high SDI regions. Conversely, low SDI regions have not yet entered the high-incidence phase for lung cancer but may face risks in the future due to the expansion of tobacco markets and worsening air pollution [23]. Middle to high SDI regions (such as China and Egypt) have the fastest rising ASPR ( EAPC=3.68), indicating they are in a “risk factor outbreak phase,” necessitating caution regarding the health costs of economic transitions. Regional comparisons further reveal heterogeneity in development patterns. The differences between East Asia (ASIR=43.4 per 100,000) and High-income North America (ASIR=37.2 per 100,000) illustrate the interplay of environmental and behavioral factors: PM2.5 exposure from coal burning and industrial emissions in East Asia is a significant driver of lung cancer [24], while North America has seen a decline in incidence rates ( EAPC=−1.62) due to strict tobacco control policies in recent years [25]. In contrast, the significant improvement in Central Asia (such as Kazakhstan, ASIR EAPC=−3.23) may be attributed to the implementation of tobacco control policies following the dissolution of the Soviet Union [26]. The GBD 2021 identified tobacco use, PM2.5 pollution, occupational carcinogens, and radon exposure as major risk factors for lung cancer. This study found that tobacco remains the leading cause of lung cancer deaths globally (accounting for 61.4%), but its impact varies regionally. Eastern Europe (66.7%) and East Asia (66.6%) have the highest proportions of tobacco-attributable deaths, associated with high male smoking rates and rising female smoking rates [25]. Notably, despite declining smoking rates in high-income countries, the lung cancer burden remains high, possibly related to the cumulative effects of smoking and delayed exposure to secondhand smoke [27].The role of PM2.5 pollution is particularly pronounced in low- and middle-income countries. The lung cancer burden in East Africa (attributable proportion 40%) and South Asia (such as India, with annual PM2.5 concentrations exceeding 90 μg/m³) is closely linked to the use of solid fuels for cooking and traffic pollution [28]. Regarding occupational exposure, Switzerland’s proportion of deaths attributable to occupational carcinogens reached 33.4%, indicating a need for enhanced protections in mining and manufacturing [29]. Additionally, the high contribution of radon exposure in Central Asia (7.4%) and Eastern Europe may be related to old building materials and geological characteristics [30].

There are significant gender differences in lung cancer, with the global ASPR for men being 1.5 times greater than that of women. However, the ASPR for women in high SDI countries is increasing more rapidly (EAPC=1.41 vs. 0.25), which may be related to rising smoking rates among women and changes in social roles[31]. Although men remain the primary patients with lung cancer, the incidence among women is gradually increasing, especially in high-income countries[32]. Age-wise, lung cancer incidence increases with age, peaking in the 75-79 age group, reflecting the impact of an aging society on the lung cancer burden.

This study provides a comprehensive assessment of the global burden of lung cancer from 1990 to 2021 and predicts trends for 2040. Although the age-standardized incidence and mortality rates for lung cancer show a slow declining trend globally, the absolute number of cases and deaths has significantly increased, particularly in low- and middle-income countries. The study emphasizes the complex relationship between socioeconomic development levels and lung cancer burden, indicating that high SDI regions face a higher burden of lung cancer, while low SDI regions may face risks in the future. Policy recommendations for lung cancer prevention and control tailored to different regions will help alleviate the global burden of lung cancer and promote improvements in public health.

Predictions show that by 2040, the global lung cancer ASPR will decrease to 31.31 per 100,000, with the burden for men (40.83 per 100,000) still higher than for women (23.52 per 100,000), but the gap between genders is getting smaller. This trend is thanks to better medical care, improved control measures, and a deeper understanding of lung cancer, while the rise in lung cancer cases among women is linked to higher smoking rates, differences in hormone receptors, and increased exposure to kitchen fumes mentioned in the gender difference analysis. The global burden of lung cancer remains heavy, posing two main challenges for health systems: high-SDI countries need to optimize early screening and targeted treatment, while low- and middle-income countries should prioritize tobacco control and pollution management [33]. For instance, putting tobacco control measures in place, increasing tobacco taxes (such as South Africa’s tax increase leading to a 20% drop in smoking rates) [34], broadening smoke-free laws (such as China’s comprehensive smoking ban policy) [35], and designing smoking cessation interventions targeted at women. Environmental governance should also be promoted, including encouraging clean energy (such as India’s “Ujjwala scheme” subsidizing liquefied petroleum gas) [36] and tightening rules on industrial emissions (such as China’s “Blue Sky Defense War”) [37]. Additionally, healthcare systems should be strengthened, promoting low-dose CT scans in low- and middle-income countries (following the U.S. NLST research model) [38], and improving access to medical care in rural areas through mobile health initiatives [39]. Working together across departments is also key for controlling lung cancer, integrating lung cancer prevention and control into urban planning (such as Copenhagen’s bike lane construction reducing traffic pollution) [40], and enhancing occupational health supervision. These steps can really help reduce lung cancer cases and ease the burden.

This study has some limitations. First, there is heterogeneity in data quality because some low-income countries lack complete cancer registration systems, which might result in missing cases (for example, only 30% of lung cancer cases are recorded in Africa). Second, the model might have some biases in its assumptions; the GBD study relies on statistical models to fill data gaps, which may underestimate the impact of genetic factors (such as the EGFR mutation subtype of lung cancer) and environmental interactions.

Third, COVID-19 may have long-term effects; this study only covers data up to 2021, and we need to keep an eye on how the pandemic affects lung cancer survival rates in the long run. Future research should integrate multi-omics data (genomics, exposure groups) to better understand how lung cancer works at a molecular level and make the best use of resources for prevention and control by looking at cost-effectiveness (such as increasing screening frequency, developing targeted drugs, etc.).

## Conclusion

This study provides the latest data on lung cancer burdens around the world, by region, and in 204 countries, showing that as of 2021, the number of lung cancer cases worldwide reached 3.25 million. From the perspective of social development levels, the burden in high-SDI areas is much higher than in low-SDI areas. Densely populated areas like East Asia and Western Europe have a heavier burden of lung cancer. Among countries, Monaco has the highest rates of lung cancer cases and deaths. In terms of gender patterns, men bear a much heavier burden of lung cancer than women, but the gap is narrowing. From an environmental perspective, tobacco is undoubtedly the biggest risk factor for developing lung cancer. By 2040, we expect the burden of lung cancer to ease somewhat. These findings provide important evidence to help countries make decisions, optimize health services, and reduce the burden of lung cancer.

## Data Availability

All relevant data are within the manuscript and its Supporting Information files.

https://ghdx.healthdata.org/gbd-2021

## Acknowledgments

We thank the contributors to the “Global Burden of Diseases, Injuries, and Risk Factors Study 2021” for their valuable work. We also sincerely thank the Institute for Health Metrics and Evaluation (IHME) for providing GBD data for this study.

## Research funding

This work was supported by the National Natural Science Foundation of China (NO. 82160900), the Gansu Province joint scientific Research Fund major project(24JRRA876),2025 Gansu Province “Innovation Star” project for graduate students (2025CXZX-952),and the Gansu University of Traditional Chinese Medicine Talent Research Initiation Fund project (2023YJRC-03).

## Contributors

All authors made significant contributions to this research. Wu Jianjun and Ma Dongjing handled the concept and methodology of the study. He Shenwen and Yang Yingying were responsible for organizing and processing data. All authors took part in coming up with and designing the study, acquiring data, and analyzing and interpreting it. Ma Dongjing wrote the article or made important revisions to its content.

## Data Sharing Statement

The GBD 2021 data resources can be accessed online through the Global Health Data Exchange (GHDx) query tool (https://vizhub.healthdata.org/gbd-results/).

## References

1. Sung H, Ferlay J, Siegel RL, Laversanne M, Soerjomataram I, Jemal A, et al. Global Cancer Statistics 2020: GLOBOCAN Estimates of Incidence and Mortality Worldwide for 36 Cancers in 185 Countries. CA Cancer J Clin. 2021;71(3):209–49.

2. Hsu JC, Wei CF, Yang SC, Lin PC, Lee YC, Lu CY. Lung cancer survival and mortality in Taiwan following the initial launch of targeted therapies: an interrupted time series study. BMJ Open. 2020;10(5):e033427.

3. Yu F, Xiao R, Li X, Hu Z, Cai L, He F. Combined effects of lung disease history, environmental exposures, and family history of lung cancer to susceptibility of lung cancer in Chinese non-smokers. Respir Res. 2021;22(1):210.

4. Shin A, Oh CM, Kim BW, Woo H, Won YJ, Lee JS. Lung Cancer Epidemiology in Korea. Cancer Res Treat. 2017;49(3):616–26.

5. Guo Q, Lu Y, Liu W, Lan G, Lan T. The global, regional, and national disease burden of breast cancer attributable to tobacco from 1990 to 2019: a global burden of disease study. BMC Public Health. 2024;24(1):107.

6. Ramadan M, Alhusseini N, Samhan L, Samhan S, Abbad T. Tobacco control policies implementation and future lung cancer incidence in Saudi Arabia. A population-based study. Prev Med Rep. 2023;36:102439.

7. Lee CT. Epidemiology of Lung Cancer in Korea. Cancer Res Treat. 2002;34(1):3–5.

8. Chang NW, Lin KC, Hsu WH, Lee SC, Chan JY, Wang KY. The effect of gender on health-related quality of life and related factors in post-lobectomy lung-cancer patients. Eur J Oncol Nurs. 2015;19(3):292–300.

9. Zhou J, Xu Y, Liu J, Feng L, Yu J, Chen D. Global burden of lung cancer in 2022 and projections to 2050: Incidence and mortality estimates from GLOBOCAN. Cancer Epidemiol. 2024;93:102693.

10. Global burden of 369 diseases and injuries in 204 countries and territories, 1990-2019: a systematic analysis for the Global Burden of Disease Study 2019. Lancet. 2020;396(10258):1204–22.

11. Global, regional, and national burden of stroke and its risk factors, 1990-2019: a systematic analysis for the Global Burden of Disease Study 2019. Lancet Neurol. 2021;20(10):795–820.

12. Clegg LX, Hankey BF, Tiwari R, Feuer EJ, Edwards BK. Estimating average annual per cent change in trend analysis. Stat Med. 2009;28(29):3670–82.

13. Knoll M, Furkel J, Debus J, Abdollahi A, Karch A, Stock C. An R package for an integrated evaluation of statistical approaches to cancer incidence projection. BMC Med Res Methodol. 2020;20(1):257.

14. Li S, Chen H, Man J, Zhang T, Yin X, He Q, et al. Changing trends in the disease burden of esophageal cancer in China from 1990 to 2017 and its predicted level in 25 years. Cancer Med. 2021;10(5):1889–99.

15. Liu N, Yang DW, Wu YX, Xue WQ, Li DH, Zhang JB, et al. Burden, trends, and risk factors for breast cancer in China from 1990 to 2019 and its predictions until 2034: an up-to-date overview and comparison with those in Japan and South Korea. BMC Cancer. 2022;22(1):826.

16. Wu B, Li Y, Shi B, Zhang X, Lai Y, Cui F, et al. Temporal trends of breast cancer burden in the Western Pacific Region from 1990 to 2044: Implications from the Global Burden of Disease Study 2019. J Adv Res. 2024;59:189–99.

17. Global burden and strength of evidence for 88 risk factors in 204 countries and 811 subnational locations, 1990-2021: a systematic analysis for the Global Burden of Disease Study 2021. Lancet. 2024;403(10440):2162–203.

18. Schmeck B, Bertrams W, Lai X, Vera J. Systems Medicine for Lung Diseases: Phenotypes and Precision Medicine in Cancer, Infection, and Allergy. Methods Mol Biol. 2016;1386:119–33.

19. Hutchings H, Wang A, Grady S, Popoff A, Zhang Q, Okereke I. Influence of air quality on lung cancer in people who have never smoked. J Thorac Cardiovasc Surg. 2025;169(2):454–61.e2.

20. Liu Z, Fang C, Sun B, Liao X. Governance matters: Urban expansion, environmental regulation, and PM2.5 pollution. Sci Total Environ. 2023;876:162788.

21. Kasymjanova G, Anwar A, Cohen V, Sultanem K, Pepe C, Sakr L, et al. The Impact of COVID-19 on the Diagnosis and Treatment of Lung Cancer at a Canadian Academic Center: A Retrospective Chart Review. Curr Oncol. 2021;28(6):4247–55.

22. Rehman MZU, Rizwan M, Sohail MI, Ali S, Waris AA, Khalid H, et al. Opportunities and challenges in the remediation of metal-contaminated soils by using tobacco (Nicotiana tabacum L.): a critical review. Environ Sci Pollut Res Int. 2019;26(18):18053–70.

23. Jani CT, Kareff SA, Morgenstern-Kaplan D, Salazar AS, Hanbury G, Salciccioli JD, et al. Evolving trends in lung cancer risk factors in the ten most populous countries: an analysis of data from the 2019 Global Burden of Disease Study. EClinicalMedicine. 2025;79:103033.

24. Lelieveld J, Klingmüller K, Pozzer A, Burnett RT, Haines A, Ramanathan V. Effects of fossil fuel and total anthropogenic emission removal on public health and climate. Proc Natl Acad Sci U S A. 2019;116(15):7192–7.

25. Been JV, Laverty AA, Tsampi A, Filippidis FT. European progress in working towards a tobacco-free generation. Eur J Pediatr. 2021;180(12):3423–31.

26. Zhou T, Wang X, Zhu Q, Zhou E, Zhang J, Song F, et al. Global trends and risk factors of laryngeal cancer: a systematic analysis for the Global Burden of Disease Study (1990-2021). BMC Cancer. 2025;25(1):296.

27. Wanderley-Flores B, Pérez-Ríos M, Montes A, Santiago-Pérez MI, Varela-Lema L, Candal-Pedreira C, et al. [Attributable mortality to tobacco consumption in Brazil, 1996-2019]. Gac Sanit. 2023;37:102297.

28. Pirie K, Peto R, Reeves GK, Green J, Beral V. The 21st century hazards of smoking and benefits of stopping: a prospective study of one million women in the UK. Lancet. 2013;381(9861):133–41.

29. The impact of air pollution on deaths, disease burden, and life expectancy across the states of India: the Global Burden of Disease Study 2017. Lancet Planet Health. 2019;3(1):e26–e39.

30. Bovio N, Wild P, Guseva Canu I. Lung Cancer Mortality in the Swiss Working Population: The Effect of Occupational and Non-Occupational Factors. J Occup Environ Med. 2021;63(12):1029–36.

31. Darby S, Hill D, Auvinen A, Barros-Dios JM, Baysson H, Bochicchio F, et al. Radon in homes and risk of lung cancer: collaborative analysis of individual data from 13 European case-control studies. Bmj. 2005;330(7485):223.

32. Malhotra J, Malvezzi M, Negri E, La Vecchia C, Boffetta P. Risk factors for lung cancer worldwide. Eur Respir J. 2016;48(3):889–902.

33. de Groot PM, Wu CC, Carter BW, Munden RF. The epidemiology of lung cancer. Transl Lung Cancer Res. 2018;7(3):220–33.

34. Koch SF. Quasi-experimental evidence on tobacco tax regressivity. Soc Sci Med. 2018;196:19–28.

35. Lin H, Chang C, Liu Z, Zheng Y. Subnational smoke-free laws in China. Tob Induc Dis. 2019;17:78.

36. Mani S, Jain A, Tripathi S, Gould CF. The drivers of sustained use of liquified petroleum gas in India. Nat Energy. 2020;5(6):450–7.

37. Zhang Q, Zheng Y, Tong D, Shao M, Wang S, Zhang Y, et al. Drivers of improved PM(2.5) air quality in China from 2013 to 2017. Proc Natl Acad Sci U S A. 2019;116(49):24463–9.

38. Tanner NT, Dai L, Bade BC, Gebregziabher M, Silvestri GA. Assessing the Generalizability of the National Lung Screening Trial: Comparison of Patients with Stage 1 Disease. Am J Respir Crit Care Med. 2017;196(5):602–8.

39. Firchow BA, Cornelius JC. Establishing a Student-Run Mobile Clinic in Appalachian Kentucky: A Community-Based Pilot Program to Address Health Inequities. J Appalach Health. 2025;6(4):97–104.

40. Nieuwenhuijsen MJ. Urban and transport planning pathways to carbon neutral, liveable and healthy cities; A review of the current evidence. Environ Int. 2020;140:105661.

